# An Extended COVID-19 Epidemiological Model with Vaccination and Multiple Interventions for Controlling COVID-19 Outbreaks in the UK

**DOI:** 10.1101/2021.03.10.21252748

**Authors:** Shuhao Zhang, Gaoshan Bi, Xiang Wang, Yun Yang, Jun Qi, Shujun Li, Xuxin Mao, Ruoling Peng, Po Yang

## Abstract

For controlling the first wave of the UK COVID-19 pandemic in 2020, a plethora of hypothetical COVID-19 models has been developed for simulating how diseases spread under different non-pharmaceutical interventions like suppression and mitigation and providing useful guidance to UK policymakers. While many models demonstrate their effectiveness on predicting and controlling the spread of COVID-19, they rarely consider consequence of incorporating the effects of potential SARS-CoV-2 variants and implementing vaccine interventions in large-scale. By December 2020, the second wave in the UK appeared to be much more aggressive with many more cases as one potentially more contagious SARS-CoV-2 variant was detected in the UK since September 2020. Meanwhile, UK has begun their first mass vaccination campaign on 8 December 2020, where three vaccines were in use including Pfizer, BioNTech and Moderna. Thus, these new issues pose an emergent need to build up advanced models for accessing effectiveness of taking both vaccination and multiple interventions for controlling COVID-19 outbreaks and balancing healthcare demands. Targeting at this problem, we conducted a feasibility study by defining a new mathematical model SEMCVRD (Susceptible [S], Exposed [E] (infected but asymptomatic), Mild [M] and Critical [C] (mild cases, severe and critical cases), [V] (vaccinated), Recovered [R] and Deceased [D]), containing two importantly new features: the combined infection of the mutant strain and the original strain and the addition of a new group who have been vaccinated. The model was fitted and evaluated with a public COVID-19 dataset including daily new infections, new deaths and daily vaccination in the UK from February 2020 to February 2021. Based on the simulation results, 1) we find under the assumption that the vaccine is equivalently effective against both the original strain and new variants of COVID-19, if the UK government implements insensitive suppression intervention for 13 weeks, COVID-19 epidemic will be controlled by the first week of April 2021 and nearly ended by the first week of May 2021. It shows that taking both vaccine and suppression interventions can effectively inhibit the spread and infection of the new mutant virus. 2) we suggest implementing a 3-weeks phased and progressive lifting intervention strategy up to a low intensity mitigation level for effectively controlling COVID-19 outbreaks in the UK. By implementing this strategy, the total number of infections in the UK will be limited to 4.2 million and the total number of deaths in the UK is 135 thousand, by the end of June 2021. The epidemic will nearly end in the early of June 2021, and the UK will not experience a shortage of medical resources. 3) On the assumption that UK has a capability of providing 600 thousand vaccinations every day, a 3-weeks phased and progressive lifting intervention strategy up to a moderate intensity mitigation level can end the epidemic by the end of May 2021. This strategy would reduce the overall infections and deaths of COVID-19 outbreaks, and balance healthcare demand in the UK.

## 1 INTRODUCTION

As of 31^st^ Jan 2020, the ongoing global epidemic outbreak of coronavirus disease 2019 (COVID-19) has spread to at least 146 countries and territories on 6 continents, resulting in 103 million confirmed cases and over 2.2 thousand deaths. ^1-2^ During the outbreak of this epidemic, empirical studies and mathematical models have shown that non-pharmaceutical interventions like suppression, mitigation or multiple interventions are effective in reducing the COVID-19 case incidence. ^3-8^ Specifically, existing hypothetical COVID-19 models include: 1) epidemic compartmental models such as the susceptible-infected-recovered (SIR) models that are easy to compute for predicting the total number of infections but likely oversimplifies complex disease processes; ^3-5^ 2) Multi-agent based models like COMOKIT, AceMod, that are able to accurately simulate the state and progress of each individual in multi-scale regions using many dynamic parameters but usually requires intensive computation resources; ^6-8^ 3) Machine learning based models that were trained using COVID-19 infection or death figures from multiple countries, but greatly relied on quality and reliability of datasets. ^9-11^ While these models demonstrate their effectiveness on predicting and controlling the spread of COVID-19 in 2020, one significant demerit is that they rarely consider consequence of incorporating the effects of potential SARS-CoV-2 variants and implementing large-scale pharmaceutical interventions. Eventually, pharmaceutical interventions are highly desirable given the socioeconomic costs of lockdowns and physical distancing. Thus, these new issues pose an emergent need to build up advanced models for accessing effectiveness of taking both vaccination and multiple interventions for controlling COVID-19 outbreaks and balancing healthcare demands.

According to Public Health England’s reports, by December 2020, the second wave in the UK appeared to be much more aggressive with many more cases as one potentially more contagious SARS-CoV-2 variant was detected in the UK since September 2020.^12^ As of 20^th^ December 2020, the regions in England with the largest numbers of confirmed cases of the variant are London, the South East and the East of England. The spreading advantage of the mutant strain relative to the original strain is quantified as: as the cumulative increase of R, the range is between 0.4 and 0.7, or as the multiplier of R, the range is between 50% and 75%.^12^ The above figures show that the infection rate of each strain in the combined infection is greatly different so that it will lead inaccurate predicted outcomes when utilising existing COVID-19 models to simulate their progression.

Meanwhile, UK has begun their first mass vaccination campaign on 8 December 2020, where three vaccines were in use including Pfizer, BioNTech and Moderna. When vaccinees becomes available, we will face a different epidemiological landscape from the early pandemic. Many populations will already have had one or more waves of COVID-19. As a result of natural immunity, the effective reproduction number will be significantly reduced from its original value in the absence of pre-existing immunity. Previously, most existing COVID-19 models only considers the effects of only implementing non-pharmaceutical interventions like suppressions or mitigations without interfering pharmaceutical interventions. In order to better understand and simulate vaccine interventions, multiple unique challenges need to be considered: the primary issue is unclear vaccine manufacturing capacity and supply for COVID-19 vaccines from Pfizer, BioNTech and Moderna. It is uncertain that when the UK’s COVID-19 vaccination programme to cover all populations; the second issue is varied vaccine effectiveness of different COVID-19 vaccines. Taking the vaccine developed by Pfizer-BioNTech as an example, it is not possible to achieve complete immunity to the COVID-19 virus after the first shot of the vaccine. According to the study, in the interval between the first and second doses, the observed vaccine efficacy against Covid-19 was 52% and that for the first 7 days after dose 2 was 91%, while reaching full efficacy against disease with onset at least 7 days after dose 2. Therefore, it is necessary to consider multiple attributes of COVID-10 vaccines affecting the future of COVID-19 control in complex non-linear ways.^13-15^

In response to above issues, a feasibility study was conducted to explore a series of epidemiological situations by adopting a hybrid of vaccine and social measure interventions for controlling the COVID-19 epidemic in the UK.^3-4^ We defined a new mathematical model SEMCVRD (Susceptible [S], Exposed [E] (infected but asymptomatic), Mild [M] and Critical [C] (mild cases, severe and critical cases), [V] (vaccinated), Recovered [R] and Deceased [D]), containing two importantly new features: the combined infection of the mutant strain and the original strain and the addition of a new group who have been vaccinated. The model was fitted and evaluated with a public COVID-19 dataset including daily new infections, new deaths and daily vaccination in the UK from February 2020 to February 2021. Based on the simulation results, 1) we find under the assumption that the vaccine is equivalently effective against both the original strain and new variants of COVID-19, if the UK government implements insensitive suppression intervention for 13 weeks, COVID-19 epidemic will be controlled by the first week of April 2021 and nearly ended by the first week of May 2021. It shows that taking both vaccine and suppression interventions can effectively inhibit the spread and infection of the new mutant virus. 2) we suggest implementing a 3-weeks phased and progressive lifting intervention strategy up to a low intensity mitigation level for effectively controlling COVID-19 outbreaks in the UK. By implementing this strategy, the total number of infections in the UK will be limited to 4.2 million and the total number of deaths in the UK is 135 thousand, by the end of June 2021. The epidemic will nearly end in the early of June 2021, and the UK will not experience a shortage of medical resources. 3) On the assumption that UK has a capability of providing 600 thousand vaccinations every day, a 3-weeks phased and progressive lifting intervention strategy up to a moderate intensity mitigation level can end the epidemic by the end of May 2021. This strategy would reduce the overall infections and deaths of COVID-19 outbreaks, and balance healthcare demand in the UK. Finally, we use scenario and sensitivity analyses to discuss how strategy effectiveness responds to possible changes in the social-epidemiological landscape that could occur before and after vaccines become available.

The remainder of this paper is arranged as follows. The model is introduced in Section 2. In the Section 3, the materials and implementation of experiment are reported. Section 4 provides detailed experimental evaluation and discussion. The conclusion and future directions are given in Section 5.

## 2 METHODOLOGY

### 2.1 Model structure

We implemented a modified SEIR model, which called SECMVRD model to account for a dynamic Susceptible [S], Exposed [E] (infected but asymptomatic), Infectious [I] (infected and symptomatic),[V] (vaccinated), Recovered [R] and Deceased [D] population state.^16-19^ For estimating healthcare needs, the infectious group was categorized into two sub-cases: Mild [M] and Critical [C] (mild cases did not require hospital beds and critical cases need hospital beds but possibly cannot get it due to shortage of health sources).^20-22^ Meanwhile, the vaccinated population was divided the into two sub-cases: Vaccinated1 [V1] and Vaccinated2 [V1], where V1 is the population who has received the first dose of vaccine, and V2 is the population who has received the second dose of vaccine.^4^ Conceptually, the modified modal is shown in Figure.1. The parameters in this model are shown in Table.1.

**Figure 1.**
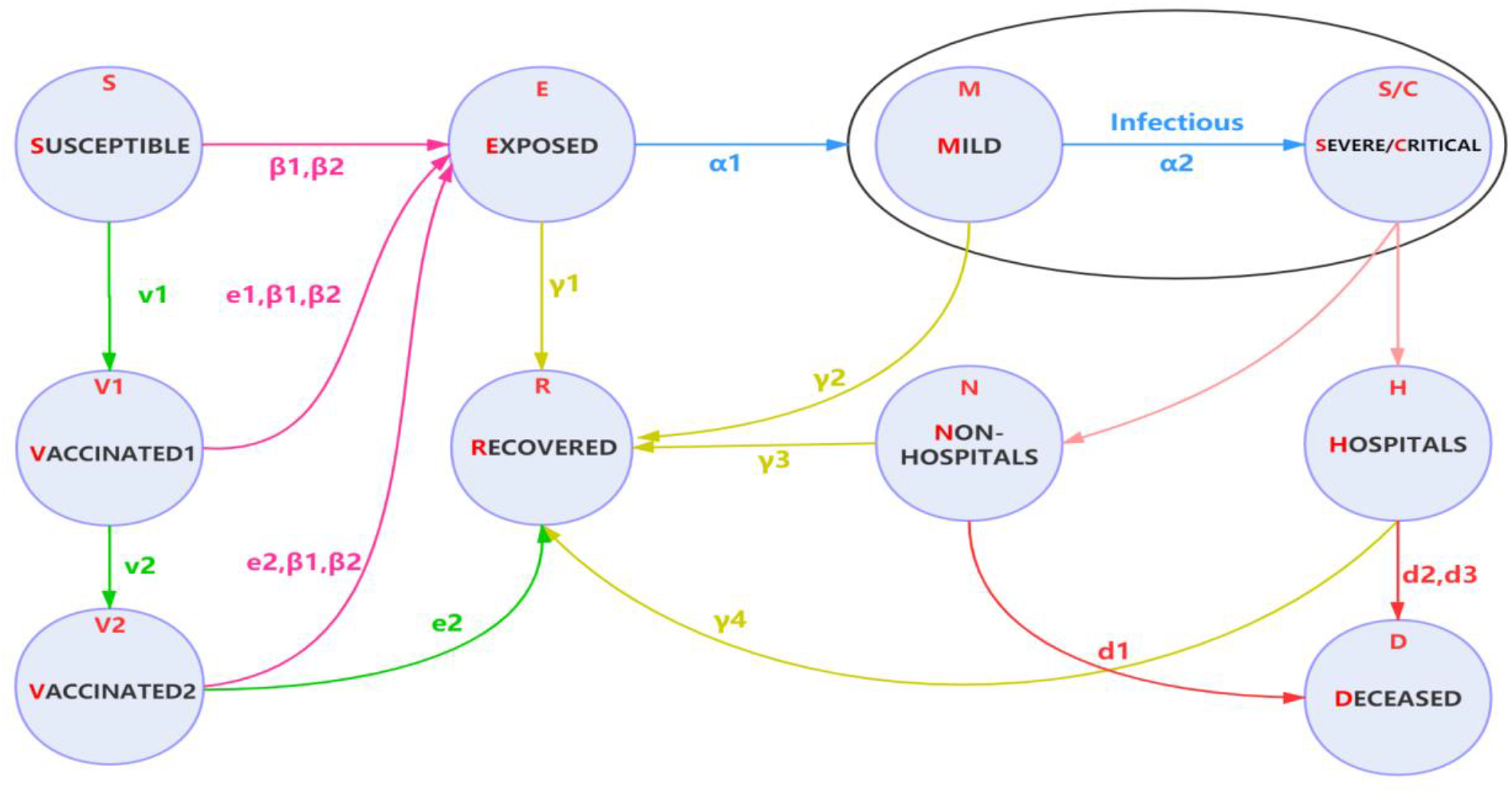
Extended SEMCVRD model structure: The population is divided into the following eight classes: susceptible, exposed (and not yet symptomatic), infectious (symptomatic), mild (mild or moderate symptom), critical (severe symptom), vaccinated, deceased and recovered (i.e., isolated, recovered, or otherwise non-infectious).

**Table 1.**
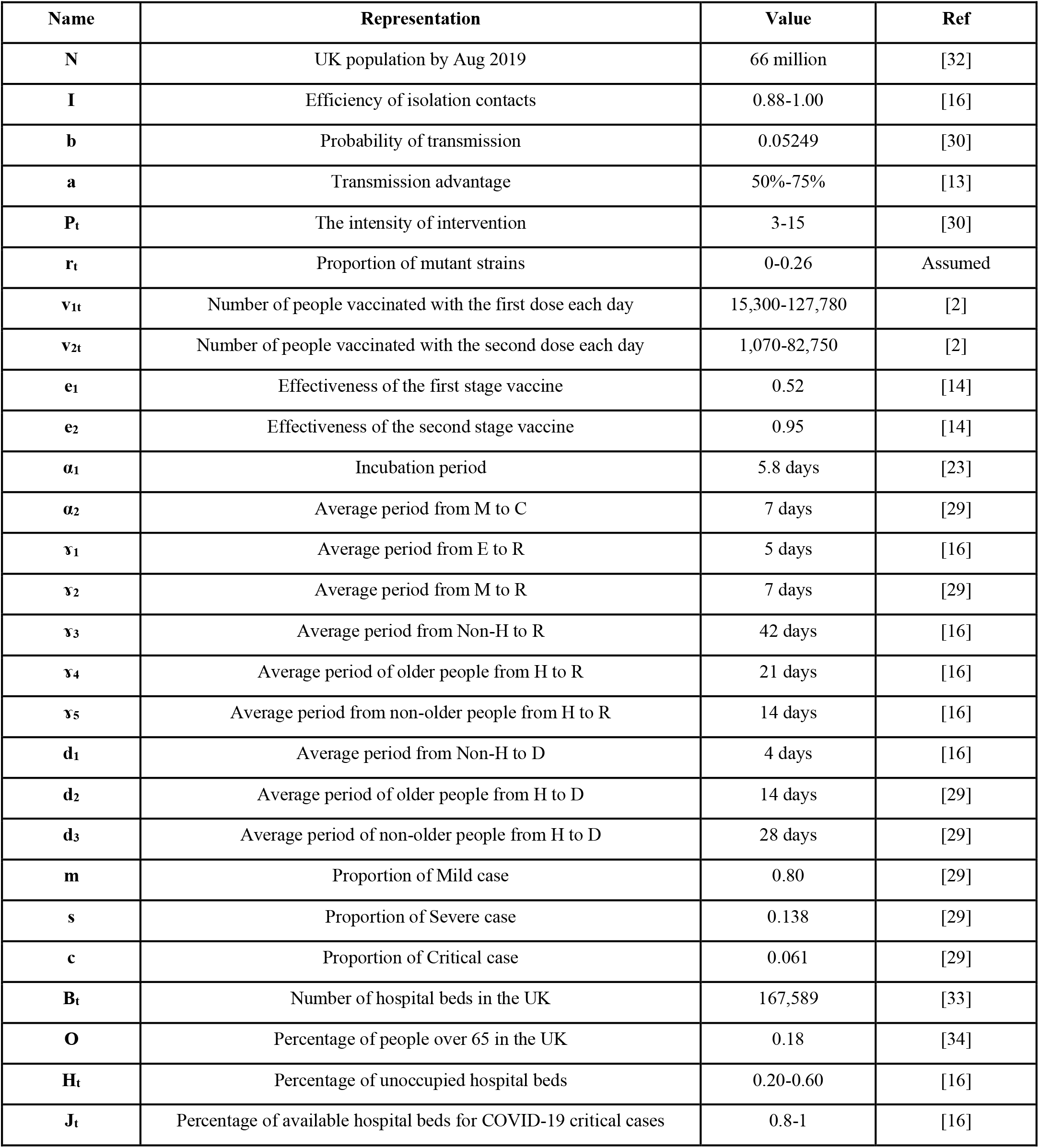
Parameters estimation in mode

The model accounted for delays in symptom onset and reporting by including compartments to reflect transitions between reporting states and disease states, where S is initial susceptible population of certain region; and incorporated an initial intervention of surveillance and isolation of cases in contain phase by a parameter β.^23-25^ If effectiveness of intervention in contain phase was not sufficiently strong, susceptible individuals may contract disease with a given rate when they come into contact with a portion of exposed population E. After an incubation period α1, the exposed individuals became the infectious population I at a ratio 1/α1.The incubation period was assumed to be 5.8 days.^26^ Once exposed to infection, infectious population started from Mild cases M to Critical cases C at a ratio a, Critical cases led to deaths at a ratio d; other infectious population finally recovered.^27-28^ It was assumed that COVID-19 can be initially detected in 2 days prior to symptom onset and persist for 7 days in mild cases and 14 days to severe cases.^29^ The susceptible population S will be vaccinated from December. The number of susceptible populations v1 will be vaccinated daily (from S to V1). 21 days after the first vaccination, V2 people receive the second vaccination every day (from V1 to V2)

In addition, one parameter was defined to measure changes in intervention intensity over time as Pt, which was presented by average number of contacts per person per day. Regarding the intensity of intervention P, it is related to the population density of an area. Our benchmark assumes that the UK without intervention is P = 15. After suppressing the intervention, P is reduced to 3.^16^ Intervention intensity was assumed within the interval [3-15], gave with a relatively accurate estimation of COVID-19 breakouts.^30^ It was assumed that transmission ratio β equals to the product of intervention intensity Pt and the probability of transmission (b) when infected. β1 and β2 respectively correspond to the infection rate of Infectious [I] and Exposed [E] (infected but asymptomatic). The value of population density and population mobility in London and the UK were calibrated and different interventions were implemented to estimate the COVID infection situation.

Notably, four important features in our model differ with other SIR or SEIR models.^24,26^ The first one was that two direct relationships were established between Exposed and Recovered population, Infections with mild symptoms and Recovered population. It was based on an observation of COVID-19 breakouts in Wuhan that a large portion (like 42.5% in Wuhan) of self-recovered population were asymptomatic or mild symptomatic.^31^ They did not go to hospital for official COVID-19 tests but actually were infected. Without considering this issue, the estimation of total infections was greatly underestimated.^20^ In order to measure portion of self-recovery population, it was assumed that exposed individuals at home recovered in 3-5 days; mild case at home recovered in 7-10 days. But if their symptoms get worse, they will be transferred to hospital.

The second feature was a consideration that the shortage of health sources (hospital beds) in the early breakouts of COVID-19 might lead to more deaths, because some severe or critical cases cannot be accommodated in time and may lead to deaths at home (non-hospital). In order to accurately quantify deaths, our model considered percentage of elder people in the UK at a ratio occupancy of available National Health System (NHS) hospital beds over time at a ratios Ht and their availability for COVID-19 critical cases at a ratio Jt. It was assumed that critical cases at non-hospital places led to deaths in 4 days; elderly people in critical condition at hospital led to deaths in 14 days and non-elderly people in critical condition at hospital led to deaths in 21 days.^30^

The third feature was to consider the outbreak of a new variant of COVID-19 in the UK in September 2020. The strain probably originated in the southeast of England in September 2020. As of 20^th^ December 2020, the areas with the most confirmed cases of this variant in England are London, the South East and the East of England. There is a consensus among all analyses that this new variant has a substantial transmission advantage over the original COVID-19 virus, with the estimated difference in the ratio of reproduction numbers varying between variant virus and original virus between 1.4 and 1.8.^13^ Based on this information, a new variant virus on the basis of the original virus was introduced to simulate the infection in the UK. The population of the region where the variant case is located accounts for about one-third of the total population of the UK. Variant confirmed cases accounted for approximately 85% of all confirmed cases in these regions. Therefore, a parameter was introduced, representing the number of cases of the mutant virus in the overall infection in the UK. The interval is roughly between 0 to 0.26 (0.85*1/3).^13^

The fourth feature was to consider the special circumstances of the UK starting vaccination in December 2020. Based on the existing vaccine information, it can be known that the vaccine is administered in two doses, so two different vaccinated populations V1 and V2 was assumed. According to statistics on the official website of the UK, the model simulates an increase in the number of two different groups of people who are vaccinated each day. In the two vaccination stages, the population has different immunity, which is represented by parameters e_1_, e_2_.^2^

### 2.2 Implementation of dynamic transmission of Model

Following previous assumptions, the implementation of dynamic transmission of our modified SEIR model follows steps as below:

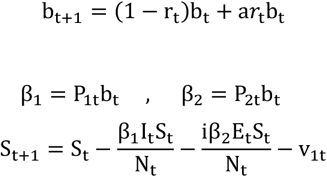

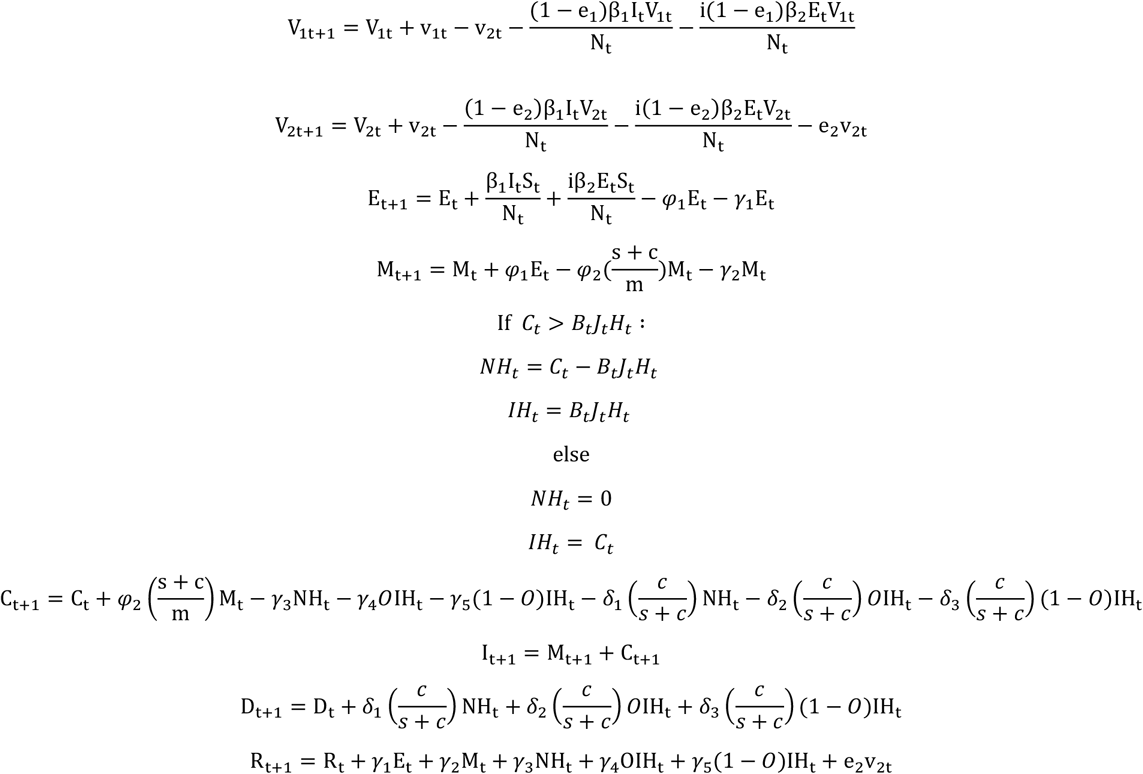

Here parameter *φ*_1_ is the transmission rate from E to M (1/ *α*_1_(incubation period)), Parameter *φ*_2_ is the transmission rate from M to C (1/ *α*_2_(average period from M to C)).

Parameter *γ*_1_ is the transmission rate from E to R (1/ γ_1_(average period from E to R)), parameter *γ*_2_ is the transmission rate from M to R (1/ γ_2_(average period from M to R)), parameter *γ*_3_ is the transmission rate from NH to R (1/ γ_3_(average period from NH to R)), parameter *γ*_4_ is the transmission rate of older people from IH to R (1/ γ_4_(average period of older people from IH to R)), parameter *γ*_5_ is the transmission rate of non-older people from IH to R (1/ γ_5_(average period of non-older people from IH to R)).

Parameter *δ*_1_ is the transmission rate from NH to R (1/ *d*_1_(average period from NH to D)), parameter *δ*_2_ is the transmission rate of older people from IH to R (1/ *d*_2_(average period of older people from IH to D)), parameter *δ*_3_ is the transmission rate of non-older people from IH to R (1/ *d*_3_(average period of non-older people from IH to D)).

## 3 EXPERIMENTS

The SEMCVRD model is explored to simulate the COVID-19 infection in the UK starting in August. Before August, the epidemic in the UK had been brought under control and stabilized. We simulated the outbreak of the second COVID-19 epidemic in the UK that started in August 2020 and included two special cases during the period: 1) At least three new variants of the virus that causes COVID-19 was detected in the UK since September 2020, where they are more infectious than the original strain. The mutant strain considered in the experimental simulation accounts for the majority of all strains of the mutant infection in the UK. 2) The official start of a COVID-19 vaccination programme in the UK began on 8^th^ December 2020. Different interventions were simulated and implemented to estimate how to control the COVID-19 epidemic after the UK lifted the lockdown on 8^th^ March 2021. A total of four types of interventions were simulated, each of which contains a different situation. The initial populations were given as UK (66.49 million). The parameter Pt representing average number of contacts per person per day was given as 15 to the UK.

### 3.1 Effectiveness of Suppression Intervention

The suppression intensity was given to reduce unaltered internal mobility of a region, where: P = 3.^35^ As shown in Figure 2, the model reproduces the temporal trend of cases observed in the UK. Starting from 1^st^ August, the current cumulative number of confirmed infections is 303,952, and the number of daily new infections is 753. By the time of the second lockdown in the UK was on 5^th^ November 2020, the cumulative number of confirmed infections at this time was 1,123,197. The number of new infections per day is 24,141. The cumulative number of infections on 5^th^ November 2020 was 3.6 times the previous number, and the number of new infections that day was 32 times higher than the previous number. The second outbreak of the COVID in the UK was undoubtedly confirmed. Observed from the time point, this outbreak was also likely to be related to a new mutant strain with a higher infection rate. After the lockdown for about one month, a clear peak can be seen in Figure 2a. The number of new infections and the number of new deaths has dropped to a certain extent. As of 2^nd^ December 2020, when the lockdown was lifted, the number of new infections per day has dropped to 14,607, which fully reflects the effectiveness of the suppression intervention measures. However, after the lift on 2^nd^ December 2020, the curve rose soon, forming the next outbreak. It can be found that the early release of intensity may increase the risk of a second outbreak. As a result, the UK implemented the lockdown strategy on 5^th^ January 2021. With the start of vaccination in mid-December, the COVID-19 epidemic was effectively contained, and the number of new infections per day also declined exponentially.

**Figure 2.**
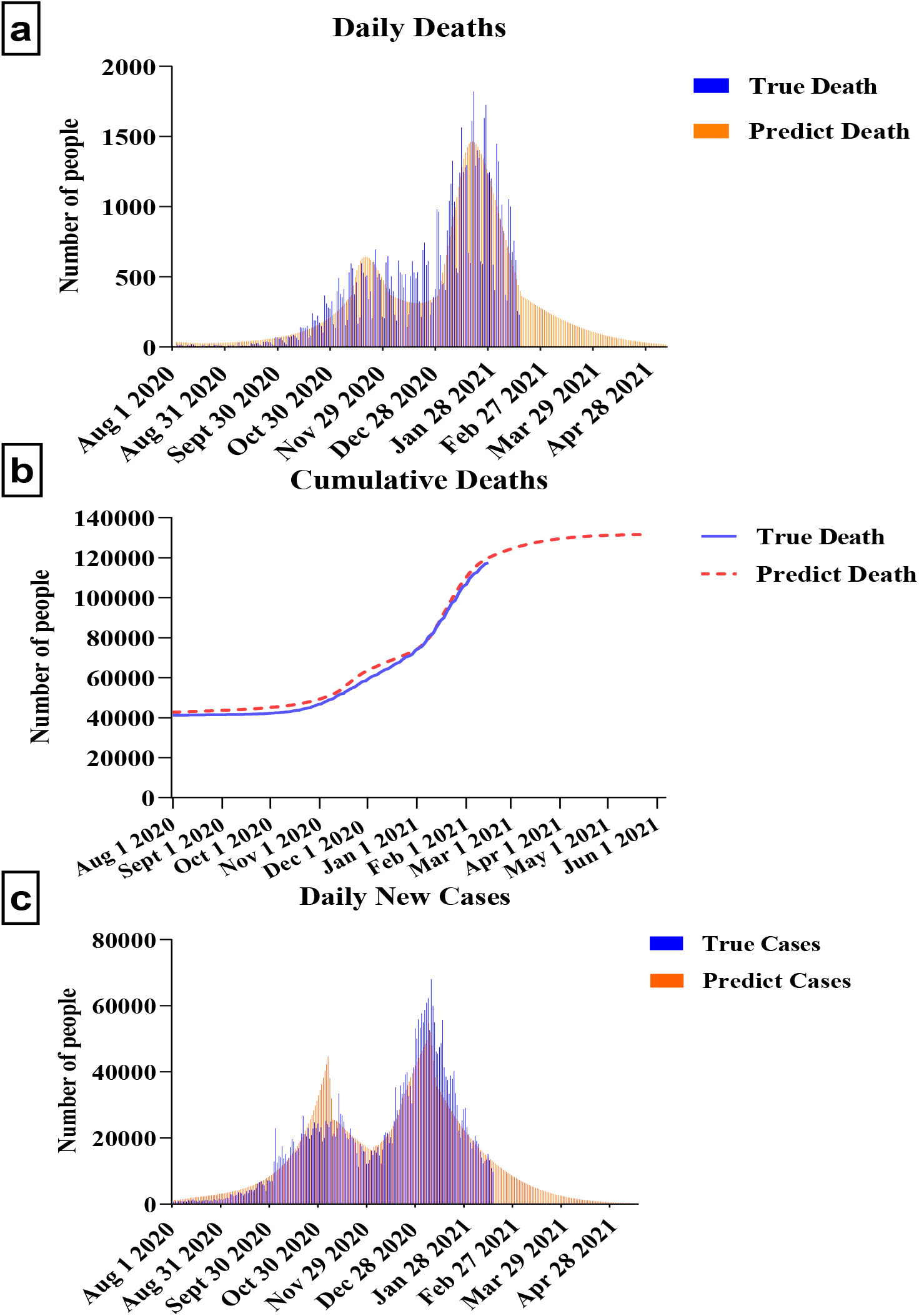
Predicting the impact of Suppression intervention on the UK after vaccination.

We estimated that the total number of infections in the UK, including exposed and infectious people, will actually reach 3,889,216, accounting for almost 0.58% of the UK population before the intensive suppression measures are lifted on 8^th^ March 2021 (the 220^th^ day). We predicted that if UK could continuously implement insensitive suppression, COVID-19 epidemic would be able to control by 9^th^ April 2021 (the 252^nd^ day) and would be nearly ended by 1^st^ May 2020 (the 274^th^ day). In this case, the total deaths by the end on 16^th^ July 2020 in the UK would be about 131,746. However, it is impossible to estimate the date when the UK will not implement any intervention restrictions under this intervention.

### 3.2 Effectiveness of Mitigation Intervention

Low-intensity, medium-intensity and high-intensity (P = 6, 8, 10) mitigation interventions in the UK on the 220^th^ day (8^th^ March 2021) were simulated when the UK lockdown was lifted, as shown in Figure 3.^29^ The simulation results showed that the mitigation strategy can delay the COVID-19 epidemic in the UK, but there will still be the next peak of the epidemic in the future, which cannot reduce the total infected population. Compared with suppression, after the mitigation measures taken by the UK on 8^th^ March 2021, the number of new infections per day has fallen more slowly, which means that the number of infections in the UK is still increasing.

**Figure 3.**
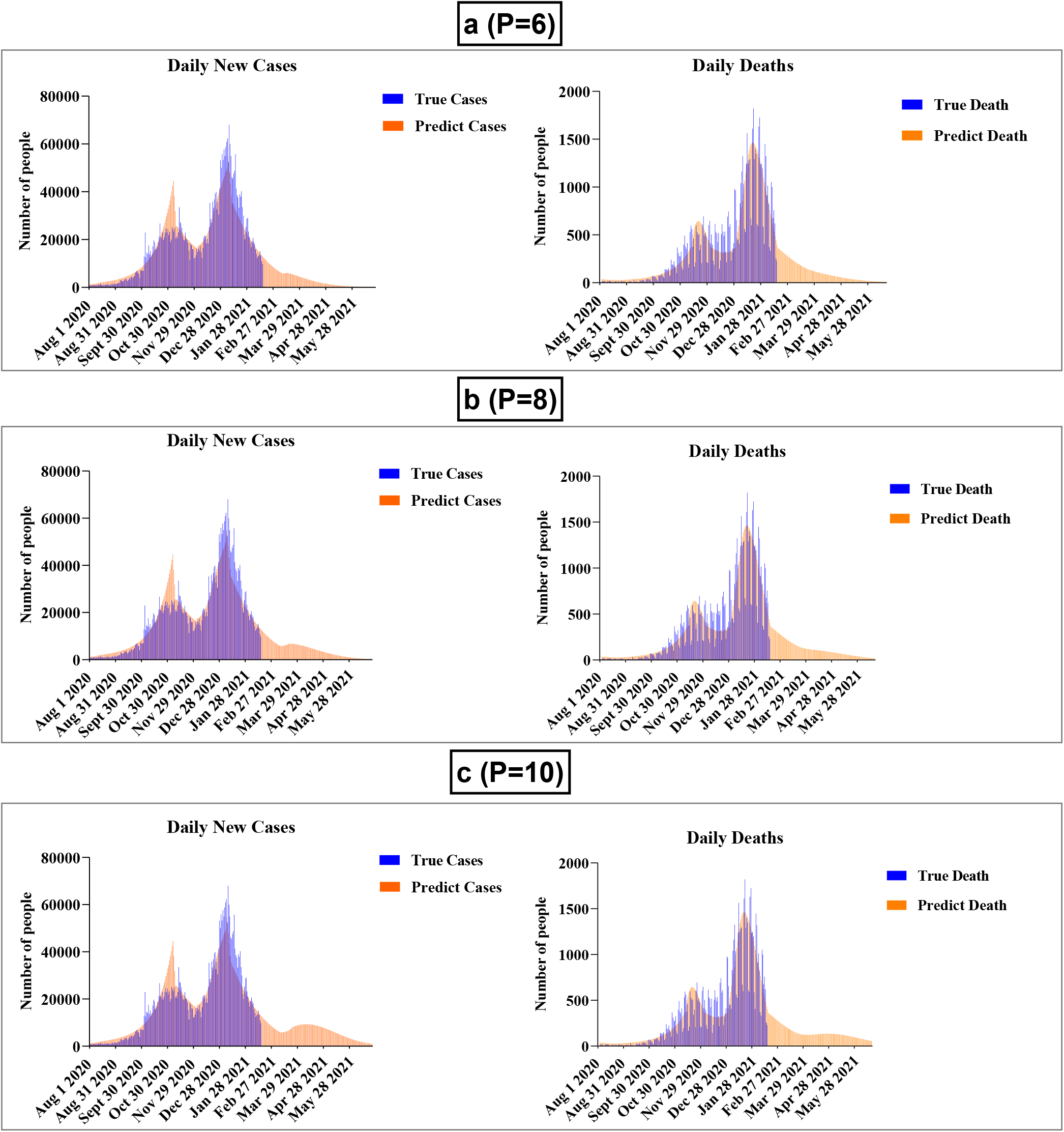
Predicting the impact of Mitigation intervention on the UK after vaccination.

The second peak of the daily infected population will reach to 6111 (M = 6), 6779 (M = 8) or 6933 (M = 10); the daily peak date of infection will be approximately on the day 227^th^ (26^th^ May 2021), 231^st^ day (8^th^ May 2021) and 251^st^ day (26^th^ April 2021). Compared with the implementation of repression, the total number of deaths in the UK will respectively increase to 133,205 (M = 6),135,131 (M = 8) or 139,604 (M = 10). The periods of COVID-19 epidemic in the UK by taking above mitigations would be extended to over 270, 292 or 318 days.

The result appeared a similar trend as findings, taking mitigation intervention in the UK enabled reducing impacts of an epidemic by flattening the curve, reducing peak incidence and overall deaths. While total infectious population may increase over a longer period, the final mortality ratio may be minimized at the end. But as same as taking suppression, mitigation need to remain in place for as much of the epidemic period as possible.

### 3.3 Effectiveness of Phase Intervention

Two possible situations were simulated in UK by implementing phase interventions as shown in Figure.4. Two possible phase intervention measures were proposed, assuming that all regions of the UK will be implemented from the 220^th^ day (8^th^ March 2021):

1. From 8^th^ March 2021, in all regions, the lockdown was gradually released by increasing the 2 number of contacts P every month.
2. From 8^th^ March 2021, in all regions, the lockdown was gradually released by increasing the 3 number of contacts P every month.

The simulation results in Figure 4.a.1 and Figure 4.b.1 show that after the lockdown was lifted, the 2-3 intensity phase intervention in the UK led to increased volatility and 2 or 3 infection peaks (the overall trend is still declining, and there is no obvious rebound) until the end of the epidemic. Compared with mitigation interventions, the total number of infections and deaths in the UK is not much different. In the phased intervention of intensity 2 and intensity 3, the total number of infections and deaths were 4,116,879 and 4,263,684, 133,192 and 135,339. The total number of deaths is about 2.3% to 6% higher than the results of the strong interventions carried out throughout the UK. Compared with suppression interventions that cannot accurately predict when the UK will not implement any intervention restrictions, premature and high-intensity opening may lead to the recurrence of epidemics, phase intervention strategies that can predict the recovery time are obviously a better choice.

**Figure 4.**
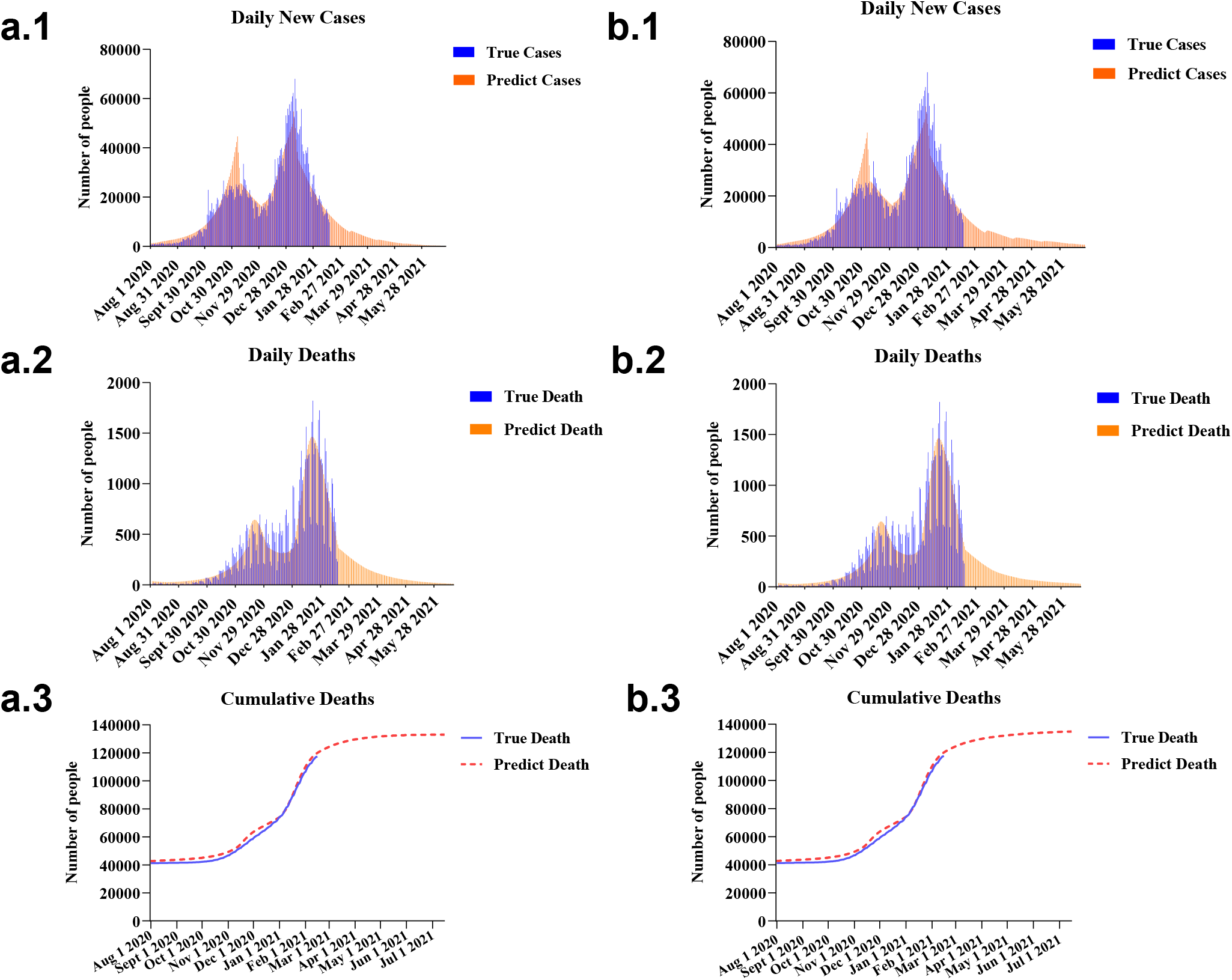
Predicting the impact of Phase intervention on the UK after vaccination.

The end time of the epidemic with intensity 2 and intensity 3 phase intervention strategies is approximately: 29^th^ April 2021 (the 272^th^ day) and 6^th^ June 2021 (the 310^th^ day), and the time for UK not implement any intervention restrictions would be about: 6 months later and 4 months later. The number of deaths and infections of the two different intensity phase intervention strategies are similar, but the intervention strategy of intensity 3 will lift all intervention restrictions in the UK two months earlier than the intervention strategy of intensity 2 (See Table 2 for details). This would be a great advantage for economic recovery.

**Table 2.** Comparison of the effects of various interventions in the UK. (TD: Total deaths (UK), TI = Total infections (UK), TN: Time for UK will not implement any intervention restrictions, E: End in half of 2021 year, TE: Approximate time for the end of the epidemic)

| Intervention | TI | TD | TN | E | TE |
| --- | --- | --- | --- | --- | --- |
| Suppression (P=3) | 4,021,223 | 131,746 | N | Y | 9<sup>th</sup> Apr 2021 |
| Lifted in March (P=6) | 4,118,092 | 133,205 | N | Y | 28<sup>th</sup> Apr 2021 |
| Lifted in March (P=8) | 4,248,430 | 135,131 | N | Y | 19<sup>th</sup> May 2021 |
| Lifted in March (P=10) | 4,542,077 | 139,604 | N | Y | 14<sup>th</sup> Jun 2021 |
| Lifted in March (P=12) | 5,331,966 | 151,300 | N | N | 22<sup>nd</sup> July 2021 |
| Lifted in March, then P+2 every month | 4,116,879 | 133,192 | 6 months | Y | 29<sup>th</sup> Apr 2021 |
| Lifted in March, then P+3 every month | 4,263,684 | 135,339 | 4 months | Y | 6<sup>th</sup> Jun 2021 |
| Lifted in March, then P+4 every month | 4,610,512 | 140,393 | 3 months | N | 25<sup>th</sup> July 2021 |
| Lifted in March, then P+5 every month | 5,229,226 | 149,436 | 2.4 months | N | 18<sup>th</sup> Aug 2021 |
| Lifted in March, then P+2 every month (Hypothetical double vaccine) | 4,051,265 | 132,207 | 6 months | Y | 14<sup>th</sup> Apr 2021 |
| Lifted in March, then P+3 every month (Hypothetical double vaccine) | 4,095,329 | 132,872 | 4 months | Y | 22<sup>nd</sup> Apr 2021 |
| Lifted in March, then P+4 every month (Hypothetical double vaccine) | 4,162,644 | 133,887 | 3 months | Y | 2<sup>nd</sup> May 2021 |
| Lifted in March, then P+5 every month (Hypothetical double vaccine) | 4,259,272 | 135,340 | 2.4 months | Y | 25<sup>th</sup> May 2021 |
| Lifted in March, then P+6 every month (Hypothetical double vaccine) | 4,377,316 | 137,133 | 2 months | Y | 8<sup>th</sup> Jun 2021 |

### 3.4 Effectiveness of Vaccine Intervention

Recommended a vaccine intervention: suppose that the number of vaccines provided for domestic use in the UK doubles after the British lockdown is lifted on 8^th^ March 2021. Combined with the previous phase intervention measures, a joint simulation is carried out to achieve an optimal intervention state. Under the circumstance that the number of vaccines available is doubled, the monthly lockdown intensity in the intervention strategy can be increased, and a phase intervention strategy with an intensity of 5 can be implemented. The experimental results are shown in Figure 5. The implementation of a phase intervention strategy with an intensity of 5 will result in increased volatility and 2 infection peaks, as shown in Figure 5.c. The overall number of infections and deaths are: 4,259,272 and 135,340. The ends of the epidemic will be around 25^th^ May 2021 (the 298^th^ day), UK will not implement any intervention restrictions at the end of May. Vaccine intervention strategies are heuristics to disengage us from focusing on social distance or the number of contacts. From another perspective, consider better interventions so that UK can lift all intervention restriction**s** earlier and the country’s economic recovery faster.

**Figure 5.**
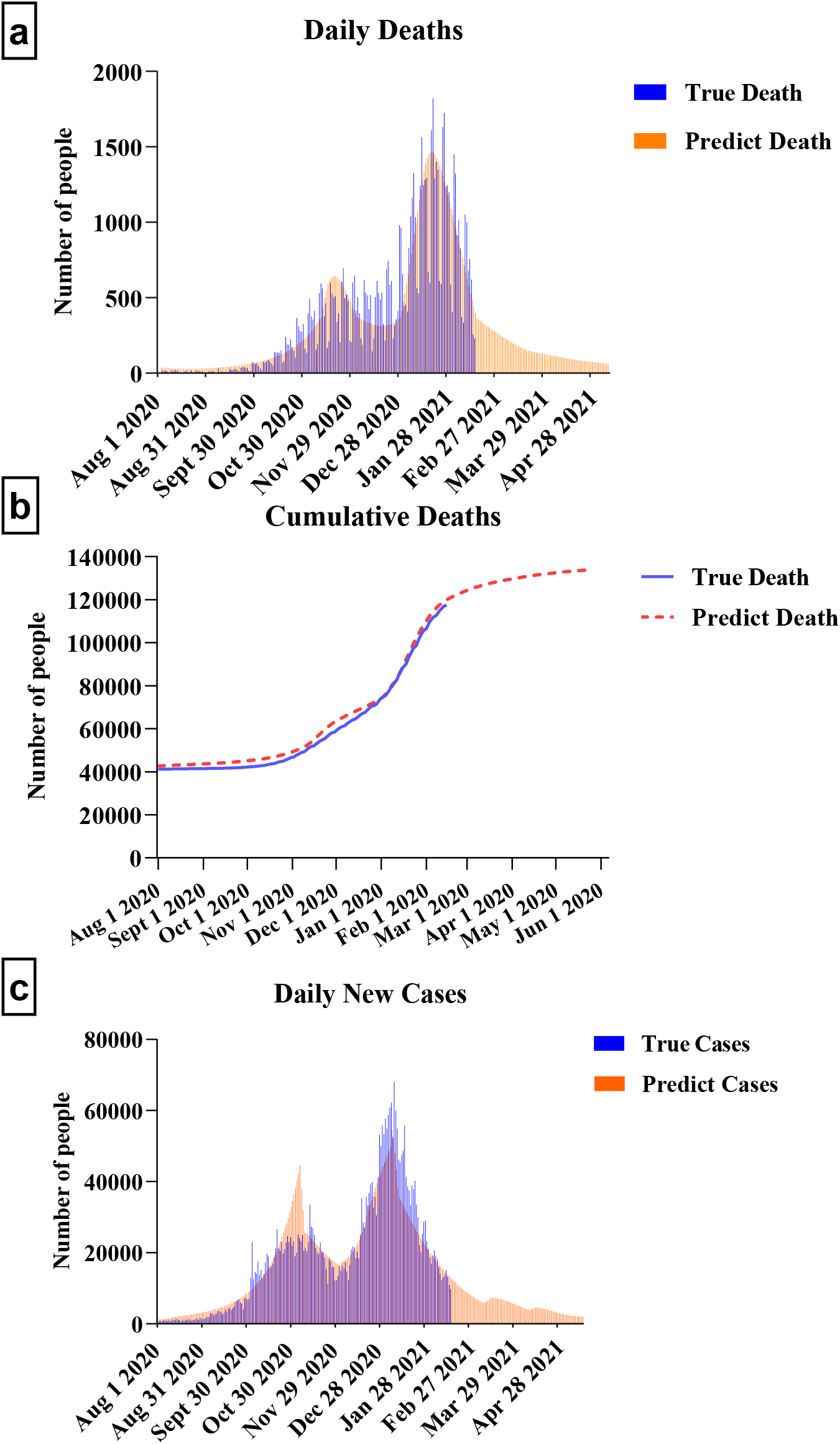
Predicting the impact of Vaccine intervention on the UK after vaccination.

### 3.5 Optimal Intervention

In addition to the intervention strategies in the previous cases, intervention strategies in other possible situations are also simulated. The initial intervention intensity is different (P = 6, 8, 10, 12) or the increased intervention intensity is different (intensity 2, 3, 4, 5 and 6), as shown in Table 2.

Firstly, the impact of different intervention intensity of mitigation intervention strategy was introduced. In mitigation intervention strategies, the choice of intervention intensity is the most important issue we need to study. In Table 2, we enumerate the number of infections and deaths under more different intervention intensities in mitigation intervention strategies. We can found from the table that when the initial intensity is greater than or equal to 10, although it can effectively delay the epidemic of COVID-19, the epidemic will obviously re-erupt by observing the curve of daily new infection number and daily new death number. When the intensity was 10 or 12, it would still cause a large number of infections and deaths. The cumulative numbers of infections and deaths of these two strategies were: 4,542,077 and 533,1966, 139,604 and 151,300, respectively, showing an exponential upward trend. Compared with mitigation intervention strategies with an intensity of 6 or 8, significantly more infections and deaths were caused. Therefore, in the selection of the intensity of the mitigation intervention strategy, we have to choose an intervention intensity below 10.

Secondly, in terms of the different increase intensities of the phase intervention strategies, we can find from Table 2 that when the monthly increase intensity is greater than 3 (the intensity is 4 or 5), it will cause the fluctuation to rise (the overall trend of the peak is upward), and there is a higher peak of infection, which makes it impossible to achieve the end of the epidemic in the first half of this year. In addition, the total number of deaths and infections of the phase intervention strategy with a monthly increase of intensity 4 or 5 are: 4,610,512 and 5,229,226, 140,393 and 149,436. Although higher intensity increments can make UK lift all intervention restriction**s** earlier, but it is not worth it if it causes a large number of infections and deaths.

Finally, different monthly lifting intensity in vaccine intervention measures were simulated. The number of infections and deaths and the end time of the epidemic are shown in Table 2. It could be found from the table that the number of infections and deaths of the stage vaccine intervention with intensity 2 or 3 were: 4,051,265, 4,095,329 and 132,207,132,872, respectively. The number of infections and deaths for strategies with an intensity of 5 or 6 were 4,259,272, 4,377,316 and 135,340,137,133, respectively. Comparing the two cases, the latter has caused an increase of about 200,000 in the number of infections, but it has not caused significantly more deaths. In addition, a strategy with an intensity of 5 or 6 can significantly shorten the time of 2-4 months, allowing the UK to lift all interventions or restrictions earlier. Therefore, if the supply of vaccines could be increased, higher-intensity phased interventions could be tried to completely lift the UK as soon as possible.

From the above, excluding the hypothesis of vaccine interventions with increasing number of vaccines, based on the balance of the overall number of deaths and infections and the time it takes for the UK to lift all intervention restrictions, it is found the use of phased interventions with increasing intensity of 3 is our best choice.

## 4 DISCUSSION

Our model suggests that when the UK began vaccination programme in December 2020, natural immunity to COVID-19 was likely to happen in many populations on account of previous infection waves. Given these likely changes to the epidemiological landscape after the vaccine becomes available, UK national suppression can be gradually lifted on the first week of March 2021, for the sake of balancing infections, deaths and potential social-economic losses due to mobility constraints. We found that between high-intensity suppression and mitigation intervention strategies, choosing a 3-weeks phased and progressive lifting intervention strategy may be an effective way to limit the total deaths but maintain essential mobility for avoiding huge economic losses in longer lockdown. Implementing this strategy can restore necessary production and commercial activities primarily, realize a slow economic recovery and ease the economic burden caused by the country’s long-term lockdown. The simulation results show that as the increasing number of people immune to the COVID virus, the epidemic would not rebound of the epidemic and normalize people’s lives as quickly as possible under the control of the lifting intensity. The total number of infections in the United Kingdom would be limited to 4.2 million; the total number of deaths in the United Kingdom would be limited to 135 thousand. UK will be able to release their social measure from suppression to mitigation with moderate intensity after four months, and the epidemic will end in early June 2020; and there will be no shortage of medical resources.

Notable, our findings also indicate that cultural difference and industrial structure of a certain country play important impacts on the implementation effectiveness of repressive interventions in controlling COVID-19 outbreaks. The success of China’s immediate suppression depends on strict restrictions on the movement of people and sufficient resources from other provinces and cities in China that have no cases of infection. Without sufficient external support, repression of the entire country will have a huge economic impact. Adopting a phased lifting intervention is appropriate. Especially, in our early work, we found that in the COVID-19 outbreak in Wuhan, a high proportion of exposed or infectious people (approximately 42%-60% of the total infected population) are self-healing. These people may think they are healthy at home because they did not go to the hospital for a COVID-19 test. An important issue is that some SEIR models predict that the number of infections in Wuhan is more than 10 times the number of confirmed cases.^36^ An excessively high-intensity release may increase the risk of another outbreak. In addition, under the assumption that the vaccine is effective against the new mutant strain, the infection of the mutant strain can be effectively suppressed under the condition of continuous vaccination, even if the infectivity of the new strain is higher than the original strain.

Lastly, there are still limitations in the simulation and analysis of our model. First of all, the prediction of our model depends on the estimation of the intensity of intervention by estimating the average number of daily contacts of infectious individuals in a certain area. Over time, each intervention will have the same or similar effects on the number of reproduction in different regions. On cultural or other issues in certain counties, the actual effect of the intensity of intervention may vary. In the UK or similar countries/regions, how to quantify the intensity of intervention requires accurate measurement of the social distance of the entire society, the isolation of cases in the family, and the family isolation of family members. Second, our model uses a variety of reasonable biological COVID-19 parameters based on the latest evidence shown in Table 1, but these assumed values may vary by population or country. For example, assuming that the average time from mild cases to severe cases is 7 days, the average time from severe cases to death of hospitalized elderly is 14 days, and so on. Changes in these variables may affect our estimates of infection and deaths in the UK. Third, our model simulates the combined infection of the original virus and the mutant virus, assuming a combined infection rate parameter, which is determined by the population of the main area infected by the mutant virus and the proportion of infection in the area. As the infection spreads, this parameter will also change. In the end, our model’s estimate of the population’s daily vaccination number is averagely estimated based on the data from the official website of the UK. In the future, with the improvement of vaccine production capacity, the parameters would be further adjusted based on real data in real time to achieve the best prediction results.

## 5 CONCLUSIONS

This paper conducted a feasibility study by defining a mathematical model called SEMCVRD, which analyzes and compares the intervention strategies to control the COVID-19 outbreak in the UK after vaccination and lifting the lockdown. The model can not only be fitted and evaluated through the public data set of the UK, but also can be fine-tuned based on the real data of other countries using the trained model to achieve prediction and analysis of other countries. The experimental results show that, in the case of continuous increase in the number of people immune to the virus, the implementation of phased and progressive lifting interventions will achieve the best balance of infection, death and economic loss. In the future, we can extend our model to achieve combined immunization of multiple vaccines and immunization strategies for people of different age groups.

## Data Availability

N/A

